# A rapid evidence map of womens health

**DOI:** 10.1101/2022.11.09.22282129

**Authors:** Deborah Edwards, Judit Csontos, Elizabeth Gillen, Ruth Lewis, Alison Cooper, Adrian Edwards

## Abstract

The rapid evidence map focuses on identifying the nature and extent of published literature on the following topic areas: healthcare professionals communication with women about womens health issues and broader health problems during clinical encounters; access to specialist healthcare; endometriosis; menopause; womens health and mental health issues, and mental health issues associated with specific conditions related to menopause or menstrual health (adenomyosis; endometriosis; fibroids; heavy menstrual bleeding, polycystic ovary syndrome and premenstrual dysphoric disorder).

The purpose of this rapid evidence map was to identify research gaps and priorities that will be beneficial to womens health in Wales. The rapid evidence map uses abbreviated systematic mapping or scoping review methods to provide a description of the nature, characteristics and volume of the available evidence.

There is a lack of primary and secondary research that explores communication between women and healthcare professionals within primary and secondary care settings. Secondary research evidence exists but there are gaps in the evidence base regarding access to services providing minor gynaecological procedures and pain management, or care for menstrual health and wellbeing, endometriosis, polycystic ovarian syndrome, menopause, heart conditions, autoimmune diseases, hypermobility spectrum disorders, myalgic encephalomyelitis, long COVID, fibromyalgia, skin conditions, or palliative and end of life care, which are priority areas identified by the Womens Health Wales Coalition (2022). There are no active funding calls exploring these topics.

Regarding endometriosis, there is a lack of review evidence regarding education and resources for health care professionals and doctors to reduce diagnostic times and improve care. There is an evidence gap for primary research regarding information, support interventions and tools for women with endometriosis to help them manage their symptoms and improve their quality of life.

A substantial amount of secondary evidence exists on menopause along with a plethora of research priorities around treatment and symptom management. It was beyond the scope of this work to determine if any research had been conducted in these priority areas since the production of the guidelines and recommendations.

There is a lack of research recommendations and review evidence that address mental health issues and specific issues that affect a womens menstrual health such as adenomyosis, fibroids, heavy menstrual bleeding and premenstrual dysphoric disorder.

**Funding statement:** The Wales Centre for Evidence Based Care was funded for this work by the Wales COVID-19 Evidence Centre, itself funded by Health and Care Research Wales on behalf of Welsh Government.

**Wales COVID-19 Evidence Centre (WCEC):** *Rapid Evidence Map: Women’s health:* Report number – REM 00045 (October 2022)

##### Rapid Evidence Map Details

##### Review conducted by

Wales Centre For Evidence Based Care

##### Review Team

▪ Deborah Edwards
▪ Judit Csontos
▪ Elizabeth Gillen

###### Review submitted to the WCEC

October 2022

###### Stakeholder consultation meeting

24^th^ October 2022

###### Rapid Evidence Map report issued by the WCEC

November 2022

##### WCEC Team

▪ Adrian Edwards, Ruth Lewis, Alison Cooper, Micaela Gal involved in drafting the topline summary, reviewing, editing, publication process.

##### This review should be cited as

REM00045. Wales COVID-19 Evidence Centre, Rapid Evidence map: Womens health. October 2022

##### Disclaimer

The views expressed in this publication are those of the authors, not necessarily Health and

Care Research Wales. The WCEC and authors of this work declare that they have no conflict of interest.

*Rapid Evidence Map: Women’s health:* Report number – REM00045 (October 2022)

**TOPLINE SUMMARY**

##### What are Rapid Evidence Maps?

Our Rapid Evidence Maps (REMs) use abbreviated **systematic mapping or scoping review methods** to provide a description of the nature, characteristics and volume of the available evidence for a particular policy domain or research question. They are mainly based on the assessment of abstracts and incorporate an a priori protocol, systematic search, screening, and minimal data extraction. They may sometimes include critical appraisal, but no evidence synthesis is conducted. Priority is given, where feasible, to studies representing robust evidence synthesis. They are designed and used primarily **to identify a substantial focus for a rapid review, and key research gaps in the evidence-base**. (*N*.*B. Evidence maps are not suitable to support evidence-informed policy development, as they do not include a synthesis of the results*.)

##### Who is this summary for?

Health and Care Research Wales

##### Background / Aim of Rapid Evidence Map (REM)

The Welsh Government Research and Development Division intends to run a commissioned funding call on understanding and tackling gender inequalities in health and social care in Wales. The purpose of this REM was to identify research gaps and priorities that will be beneficial to women’s health in Wales to inform the proposed funding call. It was decided, based on a preliminary review of the literature, feedback from an NHS public consultation exercise in Wales, and further discussion with the stakeholder group, that the REM would focus on identifying the nature and extent of the literature on the following prioritised topic areas: **healthcare professionals’ communication with women** about women’s health issues and broader health problems during clinical encounters; **access to specialist healthcare**; **endometriosis**; **menopause**; **women’s health and mental health issues, and mental health issues associated with specific conditions related to menopause or menstrual health** (adenomyosis; endometriosis; fibroids; heavy menstrual bleeding, polycystic ovary syndrome and premenstrual dysphoric disorder). Research gaps in other areas and health conditions, in which women might also experience inequality, were not explored in this REM.

##### Key Findings

###### Extent of the evidence base

▪ **Communication within health care encounters**: The evidence base included one systematic review (of endometriosis) and nine primary studies. The primary studies focused on breast cancer (n=2), maternal medicine (n=3), perinatal mental health (n=1), gynaecological conditions (n=1), and non-specific conditions (n=2). Three studies focused on specific populations: urban Africans, Iraqi Muslim refugees, and undocumented migrants. Planned and ongoing NIHR funded projects include clinicians’ perspectives of listening to women’s health, menstrual and gynaecological conditions, menopause, and women’s cancers
▪ **Access to specialist healthcare**: The evidence base consisted of 19 reviews and 9 protocols. Conditions covered were maternal medicine (n=8), sexual and reproductive health (n=5), cancer and cancer screening (n=4), perinatal mental health (n=4), mental health (n=2), HIV (n=2), and non-specific conditions (n=3). Specific populations investigated were refugees or displaced people (n=6), those in differing social, economic, and environmental circumstances (n=4), physical disabilities (n=3), homeless (n=2), migrants (n=2), experiencing intimate partner violence (n=1), and minority ethnicity black (n=1). The reviews focused on barriers and facilitators (n=10), barriers (n=5), experiences (n=3), mapping the evidence (n=3), factors (n=2), management (n=1), facilitators (n=1), predictors (n=1), associations (n=1), and prevalence (n=1).
▪ **Endometriosis**: The evidence base included 121 systematic reviews covering different topics including medical management (n=22), surgical management (n=15), biology/molecular (n=12), risk factors (n=11), and comorbid conditions (n=9). Research priorities were identified by the James Lind Alliance (JLA), NICE guideline, a Wales-specific primary study (Boivin et al 2018), and researchers within the field (n=2). Recent UK funding calls were identified covering laboratory research, aetiology of endometriosis and uterine disorders, and medical and surgical management.
▪ **Menopause**: The evidence base included 108 systematic reviews covering different topics including hormonal therapies (n=17), homeopathic therapies (n=13), non-hormonal therapies (n=10), genitourinary symptoms of menopause (n=7), alternative therapies (n=6), and lifestyle interventions (n=6). Research priorities were identified as part of a NICE guideline, by the British Menopause Society, and researchers within the field (n=3). Recent UK funding calls were identified covering reproductive and menopausal health, testosterone for the treatment of symptoms, women’s reproductive health in the workplace, and women’s health hub landscape.
▪ **Women’s health and mental health issues**: The evidence base included 37 reviews covering: perinatal mental health (n=23), general mental health (n=9), polycystic ovary syndrome (n=3), and intimate partner violence (n=2). Some reviews focused on specific populations including women in prison, women in inpatient mental health services, mental health of migrants and refugee women, and mental health of women from different minority groups. Recent UK funding calls were identified covering: young women’s mental health, women and partners who have experienced pregnancy not ending in live births, and perimenopause and the risk of psychiatric disorders.
▪ **Mental health issues associated with specific conditions related to menopause or menstrual health**: The evidence base included 10 systematic reviews covering: polycystic ovary syndrome (n=4), endometriosis (n=4) menopause (n=1), and menstruation (n=1). The reviews focused on prevalence (n=4), associations (n=4), and management (n=2).

###### Recency of the evidence base

▪ The review included evidence available (from 2012, 2018, and 2021) up until September 2022. (Separate searches were conducted for different topics, with variable time limits due to the varying volume of research published in certain areas.)

##### Summary of the evidence gaps

▪ There is a **lack of primary and secondary research** that explores **communication between women and healthcare professionals (HCPs)** within primary and secondary care settings.
▪ Secondary research evidence exists but there are **gaps in the evidence** base regarding **access to services** providing minor gynaecological procedures and pain management, or **care for menstrual health and wellbeing, endometriosis, polycystic ovarian syndrome, menopause**, heart conditions, autoimmune diseases, hypermobility spectrum disorders, myalgic encephalomyelitis, long COVID, fibromyalgia, skin conditions, or palliative and end of life care, which are priority areas identified by the Women’s Health Wales Coalition (2022). There are no active funding calls exploring these topics.
▪ Regarding endometriosis, there is a **lack of review evidence** regarding **education and resources for HCPs and doctors** to **reduce diagnostic times** and **improve care**. There is an **evidence gap** for primary research regarding **information, support interventions and tools** for women with endometriosis to help them **manage their symptoms** and improve their **quality of life**.
▪ A substantial amount of secondary evidence exists on **menopause** along with a **plethora of research priorities** around **treatment and symptom management**. It was **beyond the scope** of this REM to **determine if any research** had been conducted in **these priority areas** since the production of the guidelines and recommendations. Researchers in the field would like to see primary research conducted in the area of **quality of life**.
▪ There is a **lack of research** recommendations and review evidence that address mental health issues and specific issues that affect a women’s menstrual health such as **adenomyosis, fibroids, heavy menstrual bleeding and premenstrual dysphoric disorder**.

## 1. BACKGROUND

This Rapid Evidence Map (REM) was conducted as part of the Wales COVID-19 Evidence Centre Work Programme. The initial request to explore women’s health came from Health and Care Research Wales (HCRW) to inform the development of a proposed commissioned funding call by the Welsh Government Research and Development Division. The research team conducted a preliminary search of the evidence and presented the findings to the stakeholder group in order to identify research priorities that will be beneficial to women’s health in Wales. As a result of the stakeholder meeting and further discussion afterwards the focus for the REM was to explore the nature and extent of the literature on **healthcare professionals’ communication** with women about women’s health issues and broader health problems during clinical encounters, and their **access to specialist healthcare**. Additionally, the nature and extent of the evidence in relation to **endometriosis, menopause** and **mental health** will also be investigated as the preliminary search identified that these areas have been shown to be a priority area (Scottish Government 2021, Department of Health and Social Care 2021). The focus of the REM was also informed by the priority areas identified during an NHS public prioritisation exercise conducted in Wales and led by a member of the stakeholder group.

### 1.1 Communication within healthcare encounters

While women’s life expectancy is on average longer than men’s in the UK, evidence suggests that women experience more ill health and disability throughout the life course (Office of National Statistics 2022). The reasons for this include women’s underrepresentation in clinical trials and research (Duma et al. 2018) and that healthcare professionals’ education and the healthcare system is designed for men (Department of Health and Social Care 2022, Women’s Health Wales Coalition 2022). Moreover, communication with women in the healthcare system could also pose barriers to seeking help for certain conditions (Scottish Government 2021, Department of Health and Social Care 2021). Women often feel that healthcare professionals do not take their symptoms seriously, or they do not receive support after events, such as a miscarriage (Scottish Government 2021, Department of Health and Social Care 2021). In addition, deaf women, women with disabilities, and refugees often face further barriers to communication, as healthcare professionals often do not know sign language or translation to other languages is not directly available (Allen & Sesti 2018, British Medical Assocation 2021). The English, Scottish and Welsh Governments have all recently published or started working on Women’s Health Strategies to support women’s health and wellbeing (Scottish Government 2021, Department of Health and Social Care 2022, Welsh Government 2022). Improved communication is part of this commitment, with professional bodies recommending training on women’s health and practice-based skills, such as communication, to be part of the medical curricula (Allen & Sesti 2018, British Medical Assocation 2021). While Women’s Health Strategies are based on national consultations and surveys, little is known about women’s experiences of communication during clinical encounters throughout the life course.

### 1.2 Women’s access to specialist healthcare

In addition to communication issues the Women’s Health Wales Coalition (2022) have identified four key themes which are common reported as being of concern around women’s health. These are access to specialist services, improved data collection, support for sustainable co-production and training for health and care professionals. Access to appropriate specialist healthcare can pose an issue to many women (Women’s Health Wales Coalition 2022). Due to the development and management of health services in England and Wales, boundaries exist between different health boards and NHS Trusts which often leads to women living in one area being unable to access specialist care in another (Women’s Health Wales Coalition 2022).

### 1.3 Endometriosis

Endometriosis affects 10% of women and those assigned female at birth (Royal College of Obstretricians & Gynaecologists 2019). It occurs where cells similar to those lining the womb are found elsewhere in the body and such cells can cause inflammation, pain and the formation of scar tissue. Symptoms that are commonly reported include chronic pelvic pain, painful periods, pain during or after sex, painful urination and bowel movements, fatigue or tiredness, and difficulties getting pregnant and can vary considerably in severity from mild or no symptoms to being chronic and debilitating (Women’s Health Wales Coalition 2022). In Wales it takes an average of nine years before women receive a diagnosis (a year longer than in England) and can involve the distress of repeated medical appointments that fail to identify a cause for symptoms (Women’s Health Wales Coalition 2022). Research has shown that on average in Wales, a women will visit the doctor 26 visits to the doctor before receiving a diagnosis (Boivin et al. 2018), resulting in delays in accessing treatment. The chronic and complex nature of endometriosis requires specialist, multi-disciplinary, long-term management and the Women’s Health Wales Coalition (2022) reported that this is severely lacking across Wales with problems identified at all levels of care. NICE guidance (NICE 2017) outlines that access to specialist gynaecologists with expertise in diagnosing and managing endometriosis should be available (Boivin et al. 2018), including those sufficiently skilled and trained to undertake diagnostic laparoscopy. However, long waiting times for gynaecology appointments and surgery and a lack of access to gynaecologists with expertise in endometriosis within Wales have been reported (Women’s Health Wales Coalition 2022). With regard to mental health, anxiety and depression are commonly reported mental health outcomes (Wang et al. 2021, Delanerolle et al. 2021) along with a decreased mental and physical quality of life (Wang et al. 2021).

### 1.4 Menopause

Women and those assigned female at birth account for 52% of the population of Wales and at some point in their lives, the majority of them will experience menopause (Women’s Health Wales Coalition 2022). There is often little recognition, appreciation or support for symptoms which for some can be severely debilitating (FTWW 2019) and women are expected to carry on working regardless and may having to leave employment often requiring interventions from healthcare professionals (Women’s Health Wales Coalition 2022).

Interventions for menopausal symptoms must be considered based on the person’s circumstances, preferences, and the short- and long-term benefits and risks of treatments (NICE 2022a). Treatments, such as hormone replacement therapy (HRT), are highly effective in reducing menopausal symptoms and health risks associated with untreated menopause, including heart disease and osteoporosis (FTWW 2019). However, both primary and secondary care professionals can have limited knowledge on HRT, and they often rely on dated evidence which only focuses on the negative effects of using such treatment (FTWW 2019). This can lead to a lack of consideration for or access to timely treatment (FTWW 2019). One of the reasons for insufficient access to treatment or specialist services are the lack of education provided for both healthcare professionals (HCPs) and women about peri- menopause, menopause, and HRT throughout the life-course. Awareness about menopause and treatments could be raised in different settings, such as GP surgeries, sexual and reproductive health clinics, hospitals or screening appointments, although this has not been the case (FTWW 2019). Furthermore, people experience a lack of availability of specialist services for menopause, which could be traced back to women’s health not being a priority for funding bodies and policy (FTWW 2019).

As women’s health groups are becoming more vocal about their needs, the British Menopause Society (BMS) recommends that access to information should be provided to women about menopause, transition, and post-menopausal life to prepare them for health changes (Hamoda et al. 2020). Moreover, holistic assessment of the individual person going through menopause should be conducted, and tailored lifestyle and treatment advice, including information on HRT risks and benefits, and complimentary therapies, should be provided (Hamoda et al. 2020).

### 1.5 Mental health

Recent survey evidence has reported associations between time to diagnosis for a gynaecological health problem and mental health (BMI Healthcare 2021c, BMI Healthcare 2021b, BMI Healthcare 2021a). Delayed diagnosis if often caused by dismissal by healthcare professionals, which can originate from the lack of awareness and education on women’s health and gynaecological conditions (BMI Healthcare 2021a). Particular conditions, such as adenomyosis, polycystic ovary syndrome (PCOS) and uterine fibroids, can be disproportionately affected by long waiting times and mental health problems.

#### 1.5.1 Adenomyosis

Adenomyosis is a benign gynaecological condition caused by endometrial tissue growing in the myometrium which is the muscle layer of the womb (NICE 2013) that can affect one in 10 women in the UK (BMI Healthcare 2021a). While some people might not experience symptoms, others report heavy, painful, prolonged, and irregular menstrual bleeding, and pelvic pain (NICE 2013, BMI Healthcare 2021a). Receiving an adenomyosis diagnosis can take several years and results of a recent survey found that 42% women waited over five years for a diagnosis and 26% experienced a wait longer than 10 years (BMI Healthcare 2021a). As a result of prolonged diagnosis and living with symptoms, such as chronic pain, adenomyosis can have a severe impact on a women’s health related quality of life, mental health (anxiety and depression) and work productivity (Alcalde et al. 2021, BMI Healthcare 2021a).

#### 1.5.2 Fibroids

Fibroids (leiomyomas, polyps) are benign tumours in the myometrium, which can be varying in size and number, round and hard in appearance, consisting of smooth muscle cells and fibroblasts. Fibroids are usually symptom free, and a high number of women will develop them through the life course (NICE 2022). However, people who experience symptoms, describe back and stomach pain, painful sexual intercourse, heavy periods, constipation, and more frequent urination (BMI Healthcare 2021b). Fibroids are fairly quick to be diagnosed, with majority of people diagnosed within a year, although some people report diagnosis took over a year from the start of symptoms (BMI Healthcare 2021b). Although the incidence of depression and anxiety are lower than for other gynaecological conditions (Li et al. 2022) the impact of uterine fibroids on a women’s psychological health is still significant (BMI Healthcare 2021c, Ghant et al. 2015).

#### 1.5.3 Polycystic Ovary Syndrome

Polycystic ovary syndrome (PCOS) is an endocrine disorder, which can present as ovulation disorders, hyperandrogenism, and polycystic ovarian morphology, meaning that follicles filled with fluid are contained in the ovaries (NICE 2022b, BMI Healthcare 2021b). Getting a diagnosis has been reported to take between one to five years or even longer (BMI Healthcare 2021b). Regarding mental health, women affected by PCOS suffer from depression, anxiety (BMI Healthcare 2021b) and experience a lower quality of life compared to healthy women (Yin et al. 2021).

#### 1.5.4 Heavy menstrual bleeding

Menstrual bleeding that is heavier than normal or lasts longer than seven days is often referred to as heavy menstrual bleeding or menorrhagia (NICE 2018b). Heavy menstrual bleeding can be a symptom of fibroids and adenomyosis, among other health conditions, and can severely impact on women’s quality of life (NICE 2018a). Symptom fluctuations over the menstrual cycle in anxiety disorders, post-traumatic stress disorders and obsessive compulsive disorders have been reported by women with regular periods (Green & Graham 2022). However, there is a lack of research on how women who do not have regular periods experience mental health symptom fluctuation (Green & Graham 2022).

#### 1.5.5 Premenstrual Dysphoric Disorder

Premenstrual dysphoric disorder is distinct from the above, as its primary symptoms are psychological. Premenstrual syndrome (PMS) is the presence of psychological, physical, and behavioural symptoms between ovulation and the start of menstruation (luteal phase of menstrual cycle) (NICE 2019). Premenstrual dysphoric disorder (PMDD) is characterised as a more severe form of PMS, which results in women experiencing at least five from 11 identified psychological symptoms of PMS (NICE 2019).

## 2. FINDINGS

### 2.1 Summary of the evidence for communication within healthcare encounters

The evidence base consisted of nine primary studies (conducted in USA (n=2), UK (n=2), Italy (n=1), Sweden (n=1), Denmark (n=2), Europe (n=1) and one systematic review for women’s communication within healthcare encounters (Table 1).

**Table 1:**
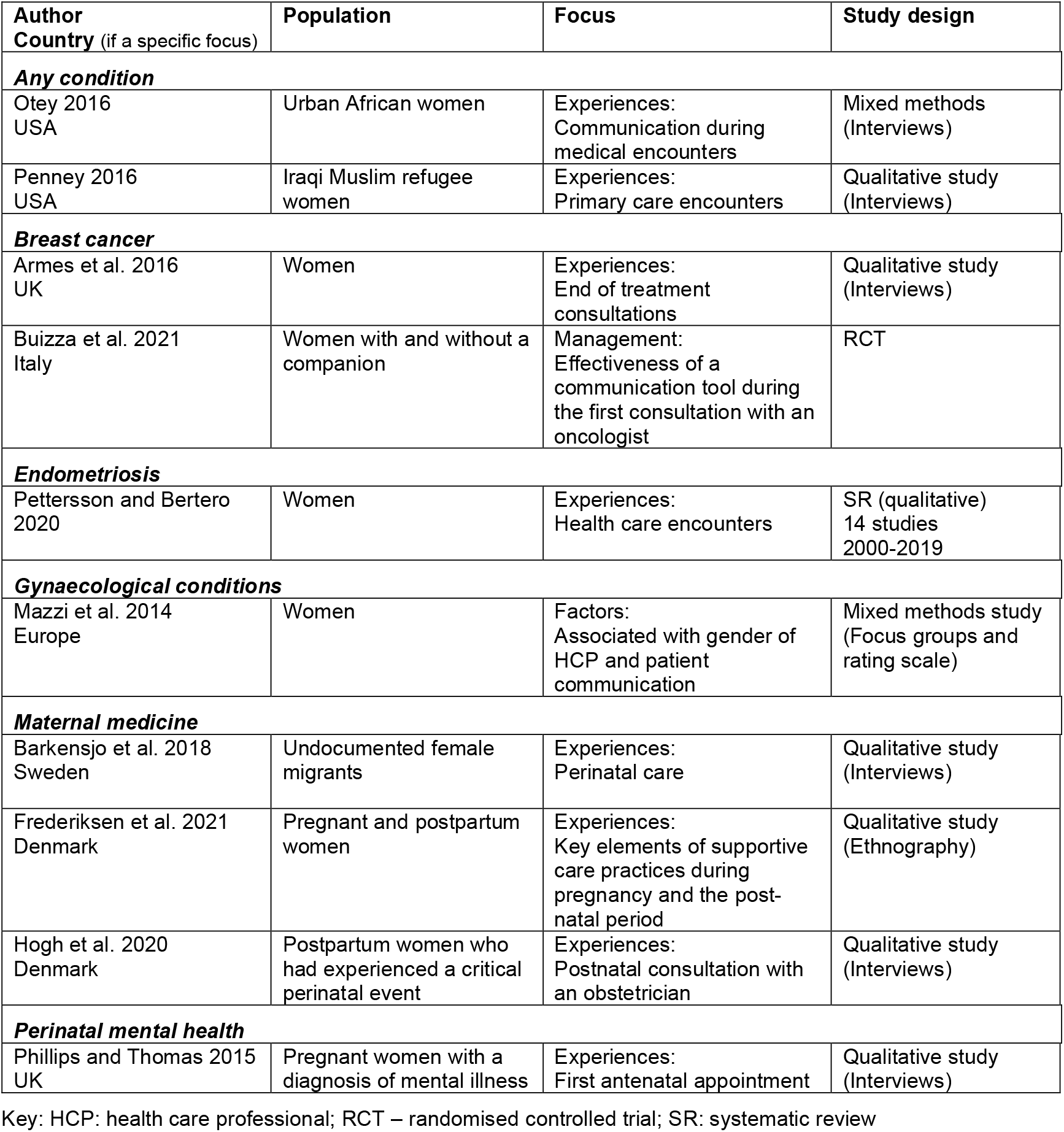
Included evidence for communication within healthcare encounters.

- The conditions covered within the primary studies were breast cancer (n=2); maternal medicine (n=3); perinatal mental health (n=1), gynaecological conditions (n=1) and any condition (n=2).
- The condition covered in the systematic review was endometriosis (n=1)
- The specific populations of women that were investigated and included urban Africans (n=1), Iraqi Muslim refugees (n=1) and undocumented migrants (n=1)
- Studies focused on experiences (n=7), management (n=1) or factors associated with gender of HCPs and patient communication (n=1).

### Planned and ongoing NIHR funded projects

Clinician’s perspectives of listening to women’s health, menstrual and gynaecological conditions (such as polycystic ovary syndrome (PCOS)), menopause, and women’s cancers.

#### 2.1.1 Bottom line summary

There is a lack of primary and secondary research that explores or addresses communication between women and healthcare professionals within primary and secondary care settings with the exception of women with endometriosis and their experiences of healthcare encounters. However, funded NIHR research focusing on HCPs’ perspectives on communication with women, who have women’s health, menstrual and gynaecological conditions (such as polycystic ovary syndrome (PCOS)), menopause, or cancer, are planned or ongoing.

### 2.2 Summary of the evidence for access to specialist healthcare

The evidence base consisted of systematic reviews (n=17); systematic review protocols (n=4); scoping reviews (n=2); scoping review protocols (n=5) that focused on women’s access to specialist healthcare (Table 2).

**Table 2:** Included evidence for women’s access to healthcare.

| **Author**<br>**Country** (if a specific focus) | **Population** | **Focus** | **Study design**<br>**Included studies**<br>**Dates of search** |
| --- | --- | --- | --- |
| ***Any condition*** |  |  |  |
| Allen and Vottero 2020 | Homeless women | Experiences:<br>Access to community based healthcare | SR (Qualitative)<br>24 included studies<br>Inception to 2018 |
| Angus et al. 2013<br>Canada | Women in differing social, economic, and environmental circumstances | Barriers:<br>Access to healthcare in Canadian settings | SR (Qualitative)<br>35 included studies<br>Dates of search NR |
| Whitfield et al. 2022<br>Canada | Women living on a low income | Mapping the evidence:<br>Access to health and social services | ScR (Mixed methods) Protocol<br>Dates of search NR |
| ***Cancer and cancer screening*** |  |  |  |
| Conti et al. 2022 | Women with breast cancer from different socioeconomic and geographic locations | Associations:<br>Relationship between geographical access to healthcare facilities (oncology services, mammography screening) and breast cancer outcomes | SR (Quantitative)<br>25 included studies<br>Dates of search NR |
| Karanth et al. 2019<br>USA | Women with ovarian cancer from different socioeconomic and ethnic backgrounds | Predictors:<br>Disparities in healthcare access | SR with MA<br>41 included studies<br>2000 to 2017 |
| Khan-Gates et al. 2015 | Women | Mapping the evidence:<br>Geographical access to mammography | SR (Quantitative)<br>21 included studies<br>2000 to 2013 |
| Pearson et al. 2021 | Women with secondary breast cancer HCPs | Barriers and facilitators:<br>Access and receipt of treatment to systemic anti-cancer therapies<br>Perspectives:<br>Access to systemic anti-cancer therapies | SR (Mixed methods) Protocol<br>2000 onwards |
| ***HIV*** |  |  |  |
| Kim and Martin 2022<br>USA (n=23/24) | Women experiencing IPV | Barriers:<br>Accessing HIV pre-exposure prophylaxis | SR (Mixed methods)<br>24 included studies<br>2007 to 2017 |
| O'Brien et al. 2018<br>HICs | Women | Facilitators:<br>Access to primary care in HICs | SR (Mixed methods)<br>44 included studies<br>2000 to 2017 |
| ***Mental health*** |  |  |  |
| DeSa et al. 2022<br>HICs | Refugee women | Barriers and facilitators:<br>Access to mental health services | SR (Qualitative)<br>12 included studies<br>Searched March 2020 |
| Wohler and Dantas 2017<br>Australia | Culturally and linguistically diverse immigrant women | Barriers:<br>Access to mental health services | SR<br>Further details NR |
| ***Maternal medicine*** |  |  |  |
| Blair et al. 2022<br>HICs | Pregnant women with physical disabilities | Experiences:<br>Access to maternity care | ScR (Mixed methods)<br>27 included studies<br>2000 to 2020 |
| Breckenridge et al. 2014 | Pregnant women with disabilities experiencing domestic abuse | Factors:<br>Associated with access to maternity care | SR (Not reported)<br>11 included studies<br>Dates of search NR |
| Higginbottom et al. 2019<br>UK | Immigrant pregnant women | Experiences:<br>Access to maternity care | SR (Not reported)<br>40 included studies<br>1990 to 2017 |
| Lawler et al. 2013 | Pregnant women with physical disabilities | Barriers and facilitators:<br>Access to maternity services | SR<br>Further details NR |
| Lennon et al. 2015 | Pregnant women | Management:<br>Effectiveness of interventions to improve access to and utilisation of prenatal care | SR (Quantitative)<br>Protocol<br>Further details NR |
| O'Mahony et al. 2022 | Perinatal refugee women | Barriers and facilitators:<br>Access to mental health services | SR (Qualitative)<br>Protocol |
| McGeough et al. 2020 | Pregnant homeless women | Barriers and facilitators:<br>Access to antenatal and postnatal care | SR (Qualitative)<br>Further details NR |
| Silva et al. 2022 | Black women | Prevalence:<br>Access to prenatal care during first trimester | SR with MA<br>Further details NR |
| ***Perinatal mental health*** |  |  |  |
| Markey et al. 2022<br>WHO European region | Refugee and asylum-seeking pregnant women | Barriers and facilitators:<br>Access to perinatal mental health care services | ScR (Not reported)<br>Protocol<br>Dates of search NR |
| O'Mahoney et al. 2022 | Pregnant refugee women | Barriers and facilitators:<br>Access to mental health care and social support | SR (Qualitative)<br>Protocol<br>2011 to present |
| Smith et al. 2019<br>UK | Pregnant women (including their partners and HCPs) | Barriers:<br>Access to mental health services | SR (Qualitative)<br>35 included studies<br>2007 to 2018 |
| Viveiros and Darling 2019<br>High resource settings | Pregnant women<br>Midwives | Barriers:<br>Access to perinatal mental health care in | ScR (Mixed methods)<br>26 included studies<br>Inception to 2000 |
| ***Sexual and reproductive health*** |  |  |  |
| Davidson et al. 2022 | Refugees and displaced women | Barriers and facilitators:<br>Access to preventive sexual and reproductive health care | SR (Mixed methods)<br>28 included studies<br>Inception to 2020 |
| Jarvis et al. 2020<br>Canada | Vulnerable women | Factors:<br>Determining access to primary sexual reproductive health care | ScR (Not reported)<br>Protocol<br>2008 to present |
| Nguyen 2020 | Women with physical disabilities | Barriers and facilitators:<br>Access to reproductive health care | ScR (Qualitative)<br>Protocol<br>2011 to present |
| O'Shea et al. 2020<br>OECD countries | Women | Barriers and facilitators:<br>Access to abortion services | SR (Mixed methods)<br>32 included studies<br>2001 to 2018 |
| Stirling et al. 2021<br>HICs | Resettled refugees and refugee-claimant women | Mapping the evidence:<br>Access and use of sexual and reproductive health services | ScR (Mixed methods)<br>Protocol<br>Search dates NR |

Key: HICs: High income countries; HIV: human immunodeficiency virus; IPV: intimate partner violence; MA: meta-analysis; NR: not reported; OCED; ScR: scoping review; SR: systematic review

- The conditions covered within the systematic reviews were maternal medicine (n=8); sexual and reproductive health (n=5); cancer and cancer screening (n=4); perinatal mental health (n=4), mental health (n=2) and HIV (n=2). Three systematic reviews did not focus on a specific condition but were on women’s health in general.
- The specific populations of women that were investigated included refugees or displaced people (n=6); those in differing social, economic, and environmental circumstances (n=4); physical disabilities (n=3); homeless (n=2); immigrants (n=2), experiencing IPV (n=1) and minority ethnicity black (n=1).
- Additionally, three reviews also included HCPs and/or women’s partners.
- The reviews focused on barriers and facilitators (n=10); barriers (n=5); experiences (n=3); mapping the evidence (n=3); factors (n=2); management (n=1); facilitators (n=1); predictors (n=1); associations (n=1) and prevalence (n=1).
- Fourteen reviews focused on healthcare access within a specific country or region and included HICs (n=5); Canada (n=3); UK (n=2); USA (n=2); OECD (n=1); WHO European region (n=1) and Australia (n=1).

#### 2.2.1 Bottom line summary

Substantial secondary research evidence exists on the topic of women’s access to specialist healthcare services, such as maternal medicine, sexual and reproductive health, cancer, perinatal and general mental health, and HIV. Research populations include women from a wide range of backgrounds and with different socioeconomic, health and ethnic characteristics. No secondary evidence was found regarding access to services providing minor gynecological procedures and pain management, or **care for menstrual health and wellbeing, endometriosis, polycystic ovarian syndrome, menopause**, heart conditions, autoimmune diseases, hypermobility spectrum disorders, myalgic encephalomyelitis, long COVID, fibromyalgia, skin conditions, or palliative and end of life care, which are priority areas identified by the Women’s Health Wales Coalition (2022). We did not find any funding calls that covered this topic.

### 2.3 Summary of the evidence for endometriosis

Searches retrieved **121** English language systematic reviews published between 2021 to October 2022. Figure 1 shows where the focus of the research lies with most studies being medical and surgical treatments (some reviews explored more than one area of research). Brady et al. (2020) reported that healthcare providers/scientists tend to prioritise research questions about cause/pathology or risk factors for endometriosis, diagnosis and screening, treatment, and fertility. Whereas patients and family members tend to prioritise questions about education/awareness, emotional impact, and comorbid conditions (Brady et al. 2020)

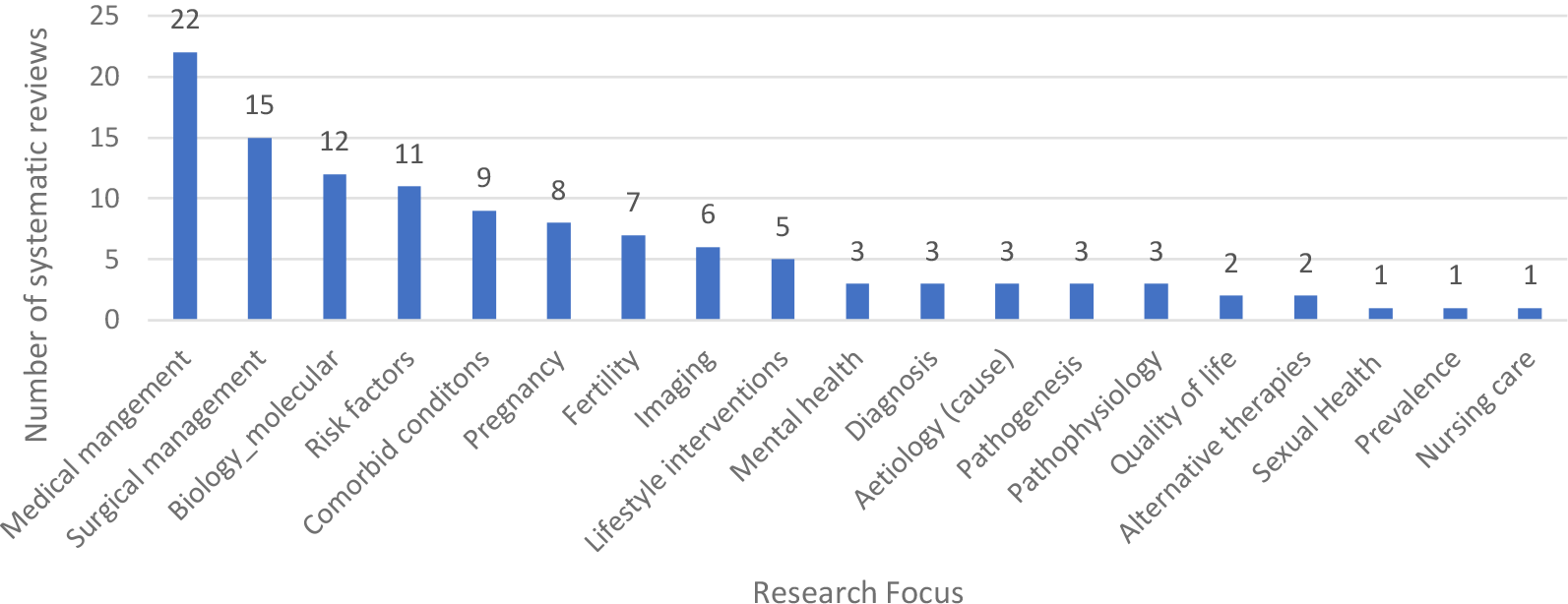
Number of systematic reviews conducted 2021 to Oct 2022 by research focus

#### Research priorities for endometriosis

- In 2017 the James Lind Alliance (JLA) published their top 10 priorities for endometriosis combining information from an online survey, systematic reviews and clinical guidelines and covered the following areas (see Appendix 1).
- The NICE guideline (NICE 2017) for endometriosis was published in 2017 and is currently being reviewed to consider whether it should be updated (DHSC 2022). The recommendations for research as set out in the current guidance can be found in Appendix 2.
- Other areas considered to be important by researchers within (As-Sanie et al. 2019, Brosens et al. 2017) and based on research already conducted in Wales (Boivin et al. 2018) are shown in Table 3.

**Table 3:** Mapping of current priority areas by research funding awards and existing systematic reviews for endometriosis.

| Priority area | Funding calls | Existing reviews |
| --- | --- | --- |
| JLA 1: Can a cure be developed for endometriosis? | Closed funding call (June 2022)<br>£250,000 in total over one to three years.<br>Scottish Government & Wellbeing of Women Funding Call for Endometriosis Research | **Not searched**<br>**Potentially evidence in this area** |
| JLA 2: What causes endometriosis? |  | **Not searched**<br>**Potentially evidence in this area** |
| JLA 3: What are the most effective ways of educating healthcare professionals throughout the healthcare system resulting in reduced time to diagnosis and improved treatment and care of women with endometriosis? |  | **No systematic reviews and no primary research studies found\*** |
| JLA 4 Is it possible to develop a non-invasive screening tool to aid the diagnosis of endometriosis? | Closed funding call (June 2022)<br>£250,000 in total over one to three years.<br>Scottish Government & Wellbeing of Women Funding Call for Endometriosis Research | **Not searched**<br>**Potentially evidence in this area** |
| JLA 5: What are the most effective ways of maximising and/or maintaining fertility in women with confirmed or suspected endometriosis? |  | Richard et al. 2021. Effect of postoperative hormonal suppression on fertility in patients with endometriosis after conservative surgery: a systematic review and meta-analysis |
| JLA 6: How can the diagnosis of endometriosis be improved?<br>Brosens et al. 2017: Better and earlier diagnosis of endometriosis |  | Simpson et al. 2021. Combating diagnostic delay of endometriosis in adolescents via educational awareness: a systematic review |
| LA 7: What is the most effective way of managing the emotional and/or psychological and/or fatigue impact of living with endometriosis (including medical, non-medical and self-management methods)? | Closed funding call (June 2022)<br>£250,000 in total over one to three years.<br>Scottish Government & Wellbeing of Women Funding Call for Endometriosis Research | **Not searched**<br>**Potentially evidence in this area** |
| JLA 8: What are the outcomes and/or success rates for surgical or medical treatments which aim to cure or treat endometriosis, rather than manage it? |  | **Previous systematic reviews all appear to be aimed at managing the condition** |
| JLA 9: What is the most effective way of stopping endometriosis progressing and/or spreading to other organs (e.g. after surgery)? |  | Chiu et al. 2022. Maintenance therapy for preventing endometrioma recurrence after |
|  |  | endometriosis resection surgery - a systematic review and network meta-analysis<br>Zakhari et al. 2021. Endometriosis recurrence following post-operative hormonal suppression: a systematic review and meta-analysis |
| JLA 10: What are the most effective non-surgical ways of managing endometriosis-related pain and/or symptoms (medical/nonmedical)? |  | **Not searched**<br>**Potentially evidence in this area** |
| NICE: Are pain management programmes a clinically and cost-effective intervention for women with endometriosis? |  | **Not searched**<br>**Potentially evidence in this area** |
| NICE: Is laparoscopic treatment (excision or ablation) of peritoneal disease in isolation effective for managing endometriosis-related pain? |  | **Not searched**<br>**Potentially evidence in this area** |
| NICE: Are specialist lifestyle interventions (diet and exercise) effective, compared with no specialist lifestyle interventions, for women with endometriosis? |  | Hanson et al. 2021. Impact of **exercise** on pain perception in women with endometriosis: A systematic review<br>Nirgianakis et al. 2022. Effectiveness of **dietary interventions** in the treatment of endometriosis: a systematic review<br>Sverrisdóttir et al. 2022. Impact of **diet** on pain perception in women with endometriosis: A systematic review<br>Tennfjord et al. 2021. Effect of **physical activity** and **exercise** on endometriosis-associated symptoms: a systematic review<br>Mardon et al. 2022. The efficacy of **self-management strategies** for females with endometriosis: a systematic review |
| NICE: What information and support interventions are effective to help women with endometriosis deal with their symptoms and improve their quality of lives? |  | **No systematic reviews and no primary research studies found\*** |
| As-Sanie et al. 2019: Quality of life |  | Kalfas et al. 2022. Psychosocial factors associated with pain and health-related quality of life in Endometriosis: A systematic review |
| As-Sanie et al. 2019: The biology of endometriosis and possible endometriosis subtypes |  | **Not searched**<br>**Potentially evidence in this area** |
| As-Sanie et al. 2019: Alternatives to hormonal therapy such as modulating angiogenesis and neuroinflammation |  | **Not searched**<br>**Potentially evidence in this area** |
| Brosens et al. 2017: Origins of endometriosis |  | **Not searched**<br>**Potentially evidence in this area** |
| <u>Boivin et al. 2018:</u> Further investigation of the care pathways for endometriosis in Wales |  | **Not searched**<br>**Potentially evidence in this area** |
| <u>Boivin et al. 2018:</u> Development of educational resources for doctors about endometriosis and young girls about menstrual health |  | **No systematic reviews and no primary research studies found\*** |
| Boivin et al. 2018: Development of support tools for women with endometriosis |  | **No systematic reviews and no primary research studies found\*** |

Key: JLA: James Lind Alliance; NICE: National Institute of Clinical Excellence
\* only searched since 2021 on one database so primary research studies may have been conducted in this area

#### Funding calls

- A funding call has just closed (June 2022) for jointly funded projects between the [Wellbeing of Women and the Scottish Government](https://www.wellbeingofwomen.org.uk/funding-opportunities/wellbeing-of-women-scottish-government-research-project-grants#:~:text=The%20total%20amount%20available%20under,with%20the%20lead%20applicant’s%20institution) in laboratory, health and translational research that aims to improve access for women to appropriate support, diagnosis and the best treatment for endometriosis.
- The National Institute of Health Research ([NIHR](https://www.fundingawards.nihr.ac.uk/)) has five active awards that focus on medical or surgical management (n=4) and management of teenagers with dysmenorrhea in primary care (n=1)
- The [Society of Endometriosis and Uterine Disorders](https://seud.org/seud-research-grant-program/) will fund aetiology, pathophysiology or treatment of endometriosis, adenomyosis, fibroids or other uterine disorders and its complications.
- The [Royal College of Obstetricians and Gynaecologists – Endometriosis Millennium Fund](https://www.rcog.org.uk/careers-and-training/awards-grants-and-prizes/endometriosis-millennium-fund/) will fund up to £5,000 in order to stimulate and encourage research (clinical or laboratory based) in the field of endometriosis.

#### Mapping of research priorities, funding calls and systematic reviews

Table 3 maps current research priorities against open and recently closed calls for funding and systematic reviews conducted in since 2021.

#### 2.3.1 Bottom line summary

A high-volume of secondary research evidence exists on the topic of endometriosis, with most systematic reviews focusing on medical and surgical treatments. Recent funding calls focus on laboratory research, aetiology of endometriosis and uterine disorders, and medical and surgical management. There is a lack of review evidence regarding education and resources for HCPs and doctors to reduce diagnostic times and improve care. Furthermore, there is an evidence gap for primary research regarding information, support interventions and tools for women with endometriosis to help them manage their symptoms and improve their quality of life.

### 2.4 Summary of the evidence for menopause

Searches retrieved **108** English language systematic reviews published between 2021 to October 2022. Figure 2 shows where the focus of the research lies with most studies being hormonal therapies, homeopathic therapies, symptom prevalence and management and non-hormonal therapies (some reviews explored more than one area of research).

**Figure 2:**
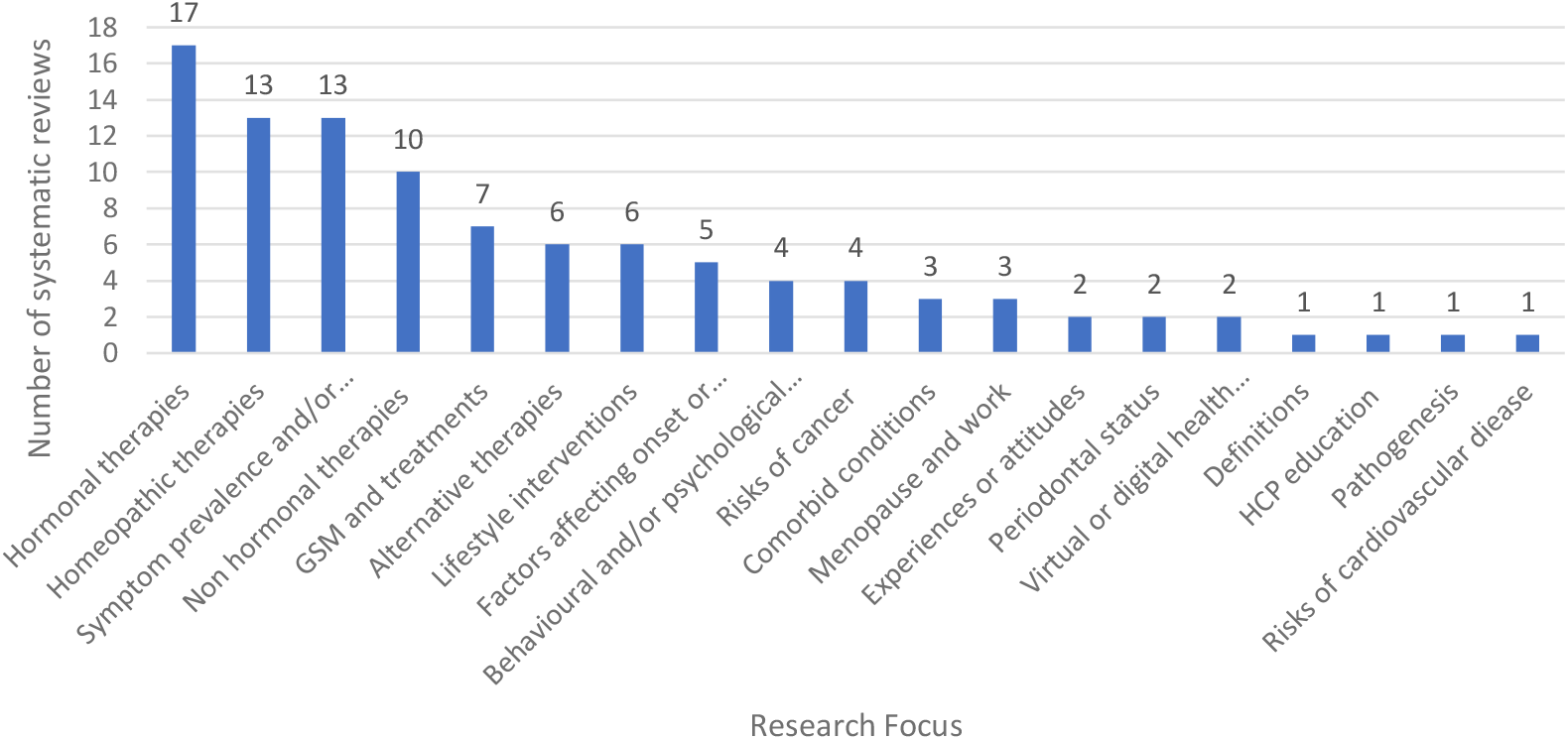
Number of systematic reviews conducted 2021 to Oct 2022 for menopause by research focus Key: GSM: genitourinary syndrome of menopause; HCP: health care professional

#### Research priorities for menopause

- The NICE guideline for menopause was published in 2015 (NICE 2015) and an update will be published in August 2023 (NICE 2022a). The recommendations for research as set out in the current guidance all focus on HRT and a broader scope that covers menopause symptoms what is being considered within the update (see Appendix 3).
- The British Menopause Society & Women’s Health Concern (Hamoda et al. 2020) have produced recommendations on hormone replacement therapy in menopausal women with breast cancer or dementia (see Appendix 4).
- Other areas considered to be important by researchers within the field (Woods & Utian 2018, El Khoudary 2017, Stuckey et al. 2020) are shown in Table 4.

**Table 4:**
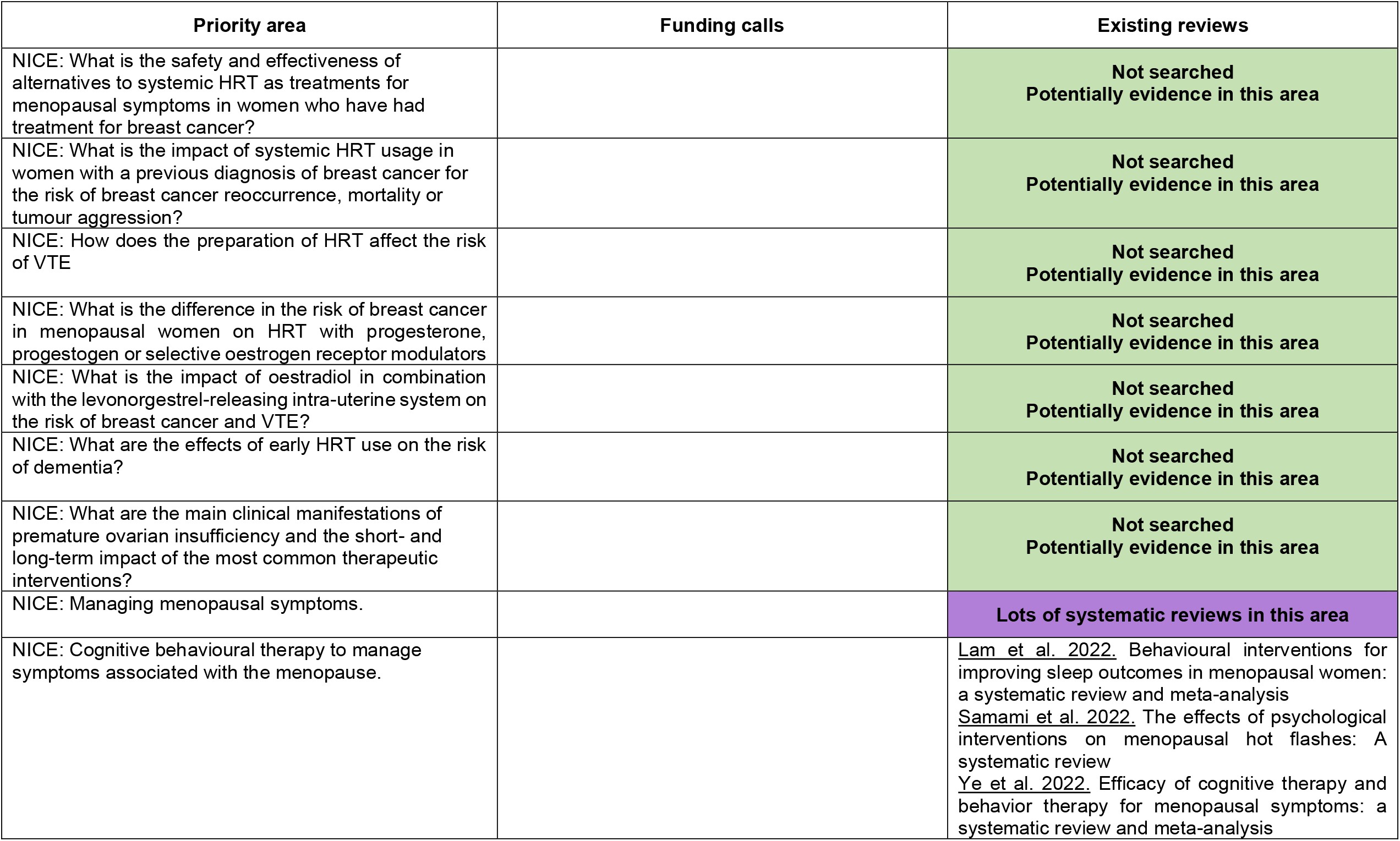
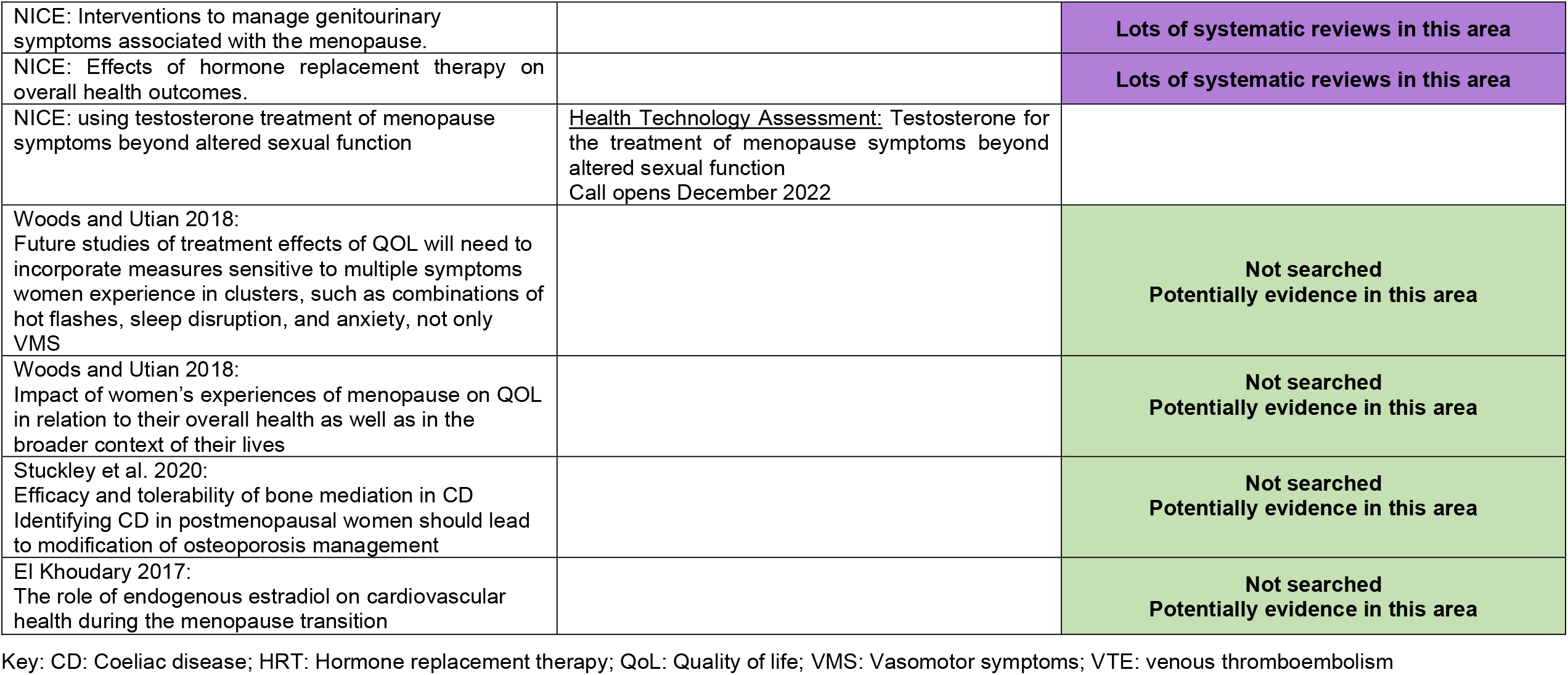
Mapping of current priority areas by research funding awards and existing systematic reviews for menopause.

#### Funding calls

- [Reproductive Health Policy Research Unit (RH PRU](https://www.nihr.ac.uk/documents/policy-research-programme-pru-reproductive-health/31173)) (closed call August 2022) which will have menstrual and menopausal health as part of its remit.
- [Health Technology Assessment:](https://www.nihr.ac.uk/funding/testosterone-for-the-treatment-of-menopause-symptoms-beyond-altered-sexual-function/31230) Testosterone for the treatment of menopause symptoms beyond altered sexual function Call opens December 2022.
- [British Menopause Society:](https://thebms.org.uk/education/british-menopause-society-education-fund/) BMS Education fund offers up to five grants will be awarded between £500 and £2,000 each year, normally to those undergoing training, but applicants not undergoing training will also be considered. Part funding or matching funding towards larger projects will be considered.
- [Department of Health & Social Care](https://www.gov.uk/government/publications/vcse-health-and-wellbeing-fund-2022-to-2025-womens-reproductive-wellbeing-in-the-workplace/vcse-health-and-wellbeing-fund-2022-to-2025-womens-reproductive-wellbeing-in-the-workplace): Health and Wellbeing Fund 2022 to 2025: women’s reproductive wellbeing in the workplace. Call closed 5^th^ August 2022.

#### Planned and ongoing NIHR funded projects

- [NIHR](https://fundingawards.nihr.ac.uk/award/NIHR135589)135589: An evaluation of the current Women’s Health Hub landscape

#### Mapping of research priorities, funding calls and systematic reviews

Table 4 maps current research priorities against open and recently closed calls for funding and systematic reviews conducted in since 2021.

#### 2.4.1 Bottom line summary

A substantial amount of secondary evidence exists on the topic of menopause, with most systematic reviews focusing on hormone therapy, complimentary or alternative therapies, symptom prevalence, genitourinary syndrome and its management, lifestyle interventions, and factors influencing onset. There are many research priorities set by NICE and the BMS and include for example further research into HRT and breast cancer or dementia, dehydroepiandrosterone (DHEA) and cancer, treatments for vasomotor symptoms (VMS). Researchers in the field would like to see research conducted into different aspects of quality of life. It was beyond the scope of this REM however, to determine if any research had been conducted in these areas since the production of the guidelines and suggestions. Current funding calls are focusing on the menopause as part of the NIHR RH PRU, testosterone treatment, and women’s reproductive wellbeing in the workplace.

### 2.5 Summary of the evidence for women’s health and mental health issues

Broad searches retrieved **37** English language systematic reviews published between 2012 to October 2022 on the topic of women’s health and mental health issues (see Table 5). The evidence base is dominated by perinatal mental health (n=23) and there are a plethora of funding calls and active grants in this area.

**Table 5:** Included evidence for women’s health and mental health issues.

| Author | Population | Study design | Focus |
| --- | --- | --- | --- |
| ***General mental health*** |  |  |  |
| Bartlett et al. 2015 | Women offenders | SR with MA | Therapeutic interventions in forensic health settings, criminal justice institutions and in the community |
| Beber 2012 | Women with intellectual disability in secure settings | NR (Not reported) | Mental health needs and development of secure services in the UK |
| Hidayati et al. 2021 | Female prisoners | ScR (Not reported) | Identifying mental health problems |
| Kapadia et al. 2017<br>UK | Pakistani women | SR (Mixed methods) | Healthcare usage rates, and the nature of Pakistani women's social networks and how they may influence mental health service use |
| Orshak et al. 2022 | Women veterans | ScR (Not reported) | Interventions of gender-sensitive care |
| Scholes et al. 2021 | Women in inpatient mental health services<br>HCPs | SR (Qualitative) | Women service users' experiences of inpatient mental health services and staff experiences of providing care |
| Wohler and Dantas 2017<br>Australia | Culturally and linguistically diverse<br>immigrant women | SR (Qualitative) | Prevalence of mental health disorders |
| Stanton et al. 2016 | Female prisoners after release from jail or<br>prison | SR (Not reported) | Understanding of the mental health issues of women released from jail or prison |
| Patten et al. 2021 | Women | SR (Quantitative) | Effectiveness of exercise interventions on mental health outcomes |
| ***Intimate partner violence*** |  |  |  |
| Paphitis et al. 2022 | Women survivors of IPV | Realist review | Management:<br>Effectiveness of psychosocial interventions to support recovery |
| Reyes et al. 2021<br>USA | Hispanic women who are victims of IPV | ScR (Mixed methods) | Mapping the evidence |
| ***Perinatal mental health*** |  |  |  |
| Ahmad and Vismara 2021 | Pregnant women | RR (Not reported) | Psychological impact of COVID-19 |
| Black et al. 2019 | Postpartum indigenous women | SR with MA | Prevalence of postpartum MH disorders |
| Coates and Foureur 2019 | Pregnant women with MH concerns | ScR (Mixed methods) | Role and competence of midwives in delivering MH care |
| Daehn et al. 2022 | Pregnant women | SR (Quantitative) | Perinatal MH literacy |
| de Oliveira et al. 2014 | Women who suffer IPV during pregnancy | SR (Quantitative) | Relationship between IPV and MH disorders |
| Demissie and Bitew 2021 | Pregnant or lactating women | SR with MA | MH effects of COVID-19 |
| Everitt et al. 2022 | Pregnant women | ScR (Not reported) | Educational innovations and teaching strategies used to build skills and knowledge in HCPs |
| Ghahremani et al. 2022 | Pregnant women | SR (Not reported) | Prevention, timely diagnosis and treatment, and referral to specialized services |
| Hansotte et al. 2017 | Low-income postpartum women in Western countries | SR (Not reported) | Factors that hinder and improve access to postpartum depression treatment in low-income women after screening |
| Kapetanovic et al. 2014 | HIV-seropositive women | SR (Not reported) | MH-related variables |
| Kesim et al. 2019 | Pregnant women | SR (Quantitative) | Effectiveness of exercise on MH |
| Lever Taylor et al. 2018 | Pregnant women partners | SR (Qualitative) | Views of perinatal MH care |
| Lucas et al. 2019<br>OECD | Pregnant women (< 20 years) | SR (Qualitative) | Perceptions of MH / wellbeing during and after pregnancy |
| Megnin-Viggars et al. 2015 | Women with (or at risk of developing) antenatal or postnatal MH problems) | SR (Qualitative) | Experience of the care for MH problems |
| Mukherjee et al. 2014 | Pregnant women in correctional facilities | SR (Quantitative) | Prevalence, determinants of MH problems |
| Nilaweera et al. 2014 | South Asian Pregnant women who have migrated to HICs | SR (Quantitative) | Prevalence, determinants of MH problems |
| Noonan et al. 2017 | Midwives | NR (Not reported) | Experiences: caring for women with MH issues |
| Owais et al. 2020 | Indigenous pregnant women | SR with MA | Prevalence of MH issues |
| Viswanathan et al. 2021 | Pregnant women with MH disorders | SR with MA | Effectiveness of pharmacotherapy |
| Waqas et al. 2022 | Pregnant women | SR with MA | Effectiveness of screening programmes for perinatal depression and anxiety |
| Watson et al. 2019 | Ethnic minority pregnant women in Europe | SR (Mixed methods) | Experiences of perinatal MH services |
| Yan and Li 2022 | Women with or without MH issues | SR with MA | Effectiveness of mindfulness-based interventions |
| Yu et al. 2022 | High-risk pregnant women | SR (Quantitative) | Effectiveness of nonpharmacological interventions |
| ***Polycystic ovary syndrome*** |  |  |  |
| Conte et al. 2015 | Women | NR (Quantitative) | Mapping the evidence on physical activity for improving MH |
| Warne et al. 2022 | Women participating in fertility treatment | SR with MA | Effectiveness of group psychological interventions in improving MH |
| Yin et al. 2021 | Women | SR with MA | Prevalence of MH problems |

Key: HPC: health care professionals; IPV: intimate partner violence; MA: meta-analysis; MH: mental health; NR: narrative review; OECD: Organisation for Economic Co-operation and Development; RR: rapid review; ScR: scoping review; SR: systematic review

There is also a significant amount of evidence for the mental health of women in general and within this the research focuses specifically on the following populations.

- Women in prison.
- Women in inpatient mental health services.
- Mental health of immigrants and refugee women.
- Mental health of women from different minority groups.

Specific searches (2018 to 2022) were conducted for the conditions related to menopause and menstrual health and wellbeing using the conditions listed in the Women’s Health Wales Coalition document and included adenomyosis; endometriosis; fibroids; heavy menstrual bleeding; PCOS, PMDD. The evidence base consisted of **ten** systematic reviews (see Table 6).

**Table 6:**
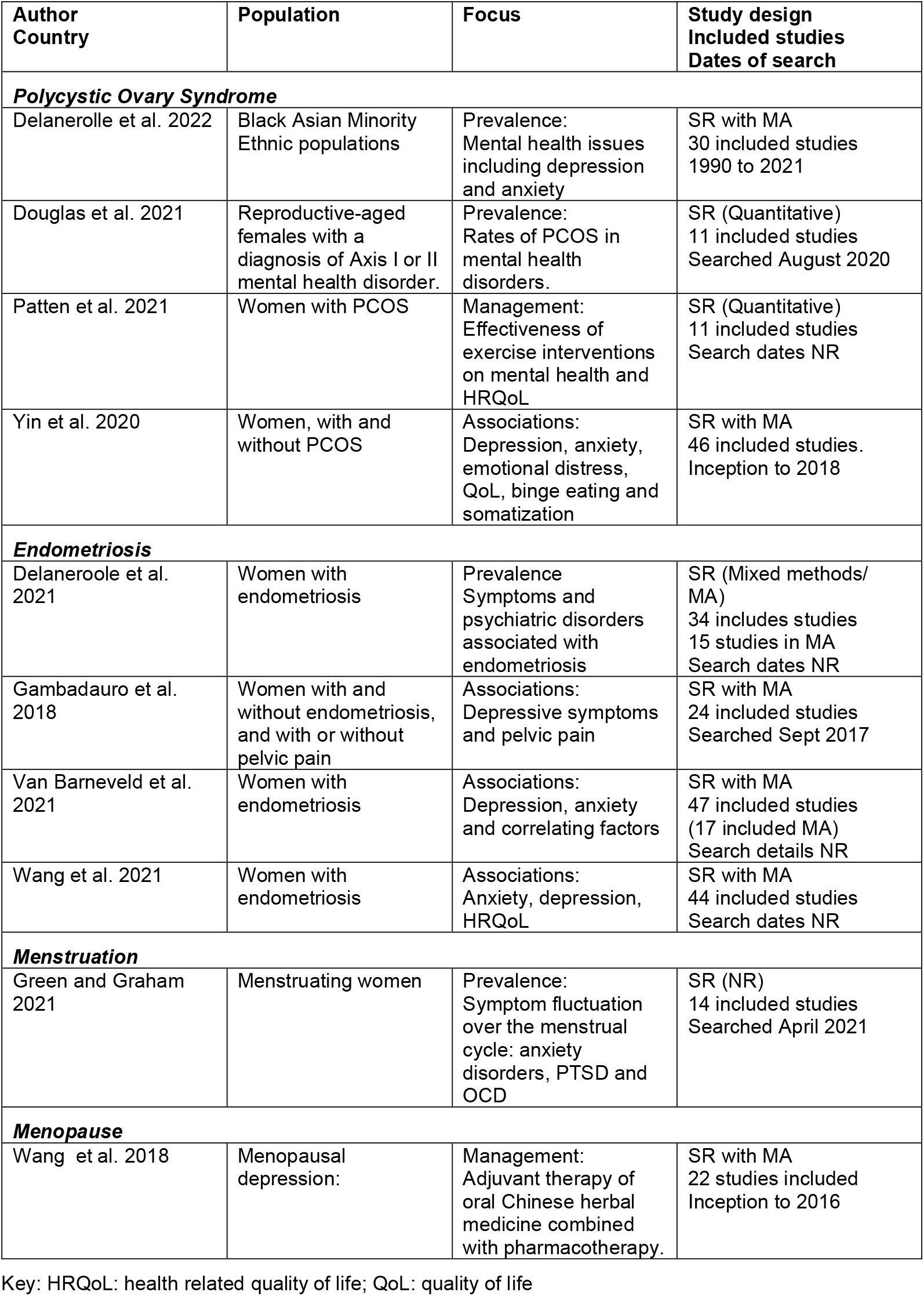
Included evidence for mental health issues across specific conditions.

| Author<br>Country | Population | Focus | Study design<br>Included studies<br>Dates of search |
| --- | --- | --- | --- |
| ***Polycystic Ovary Syndrome*** |  |  |  |
| Delanerolle et al. 2022 | Black Asian Minority<br>Ethnic populations | Prevalence:<br>Mental health issues<br>including depression<br>and anxiety | SR with MA<br>30 included studies<br>1990 to 2021 |
| Douglas et al. 2021 | Reproductive-aged<br>females with a<br>diagnosis of Axis I or II<br>mental health disorder. | Prevalence:<br>Rates of PCOS in<br>mental health<br>disorders. | SR (Quantitative)<br>11 included studies<br>Searched August 2020 |
| Patten et al. 2021 | Women with PCOS | Management:<br>Effectiveness of<br>exercise interventions<br>on mental health and<br>HRQoL | SR (Quantitative)<br>11 included studies<br>Search dates NR |
| Yin et al. 2020 | Women, with and<br>without PCOS | Associations:<br>Depression, anxiety,<br>emotional distress,<br>QoL, binge eating and<br>somatization | SR with MA<br>46 included studies.<br>Inception to 2018 |
| ***Endometriosis*** |  |  |  |
| Delaneroole et al.<br>2021 | Women with<br>endometriosis | Prevalence<br>Symptoms and<br>psychiatric disorders<br>associated with<br>endometriosis | SR (Mixed methods/<br>MA)<br>34 includes studies<br>15 studies in MA<br>Search dates NR |
| Gambadauro et al.<br>2018 | Women with and<br>without endometriosis,<br>and with or without<br>pelvic pain | Associations:<br>Depressive symptoms<br>and pelvic pain | SR with MA<br>24 included studies<br>Searched Sept 2017 |
| Van Barneveld et al.<br>2021 | Women with<br>endometriosis | Associations:<br>Depression, anxiety<br>and correlating factors | SR with MA<br>47 included studies<br>(17 included MA)<br>Search details NR |
| Wang et al. 2021 | Women with<br>endometriosis | Associations:<br>Anxiety, depression,<br>HRQoL | SR with MA<br>44 included studies<br>Search dates NR |
| ***Menstruation*** |  |  |  |
| Green and Graham<br>2021 | Menstruating women | Prevalence:<br>Symptom fluctuation<br>over the menstrual<br>cycle: anxiety<br>disorders, PTSD and<br>OCD | SR (NR)<br>14 included studies<br>Searched April 2021 |
| ***Menopause*** |  |  |  |
| Wang et al. 2018 | Menopausal<br>depression: | Management:<br>Adjuvant therapy of<br>oral Chinese herbal<br>medicine combined<br>with pharmacotherapy. | SR with MA<br>22 studies included<br>Inception to 2016 |

Key: HRQoL: health related quality of life; QoL: quality of life

- The conditions covered were PCOS (n=4), endometriosis (n=4), menstruation (n=1) and menopause (n=1).
- One review for PCOS investigated the prevalence of mental health issues in Black, Asian and minority ethnic populations.
- There was no systematic review evidence for mental health and adenomyosis; fibroids; heavy menstrual bleeding or PMDD.
- The reviews focused on prevalence (n=4), associations (n=4) and management (n=2).

#### Funding calls

- [The Pilgrim Trust:](https://www.thepilgrimtrust.org.uk/young-womens-mental-health-grants/) Young women’s mental health grants. £20,000 to £30,000 per year for a 3 year project. For 2022 the focus is North East and North West England and Northern Ireland.
- NIHR Funding call: [22/80 Interventions to promote mental health and wellbeing among young women](https://www.thepilgrimtrust.org.uk/young-womens-mental-health-grants/). What interventions are effective to promote good mental health and wellbeing among young women aged 12-24? Call opened 28 June 2022 and closes 29 November 2022.
- NIHR Funding call: [22/82 Improving mental health outcomes for women and partners who have experienced pregnancy not ending in live births](https://www.nihr.ac.uk/funding/2282-improving-mental-health-outcomes-for-women-and-partners-who-have-experienced-pregnancy-not-ending-in-live-births/30810). Which interventions are the most impactful in improving mental health outcomes in women or/and partners experiencing a pregnancy not ending in a live birth in the UK? Call opened 28 June 2022 and closes 29 November 2022.

#### Active grants

Medical Research Council: Perimenopause and risk of psychiatric disorders: a longitudinal, population-based study. Funding period: 2022-2025

https://gtr.ukri.org/project/0510802D-9354-4182-AA13-59CA915CF45A

#### Research priorities for mental health across a range of conditions

Across the condition specific NICE guidance and the National Association for Premenstrual Syndrome guidance there are no specific recommendations that focus on mental health and adenomyosis; endometriosis; menopause; fibroids; heavy menstrual bleeding; PCOS or PMDD.

#### 2.5.1 Bottom line summary for women’s health and mental health issues

A high-volume of secondary research evidence exists on the topic of perinatal mental health and general mental health issues, especially across seldom heard populations with funding calls also focusing on these areas. There is a lack of research recommendations and review evidence that address mental health issues and specific issues that affect a women’s menstrual health such as adenomyosis, fibroids, heavy menstrual bleeding and PMDD.

### 2.6 Summary tables

## 3. STRENGTHS AND LIMITATIONS OF THIS REM

A strength of this REM is that literature searches were conducted by an information specialist and an experienced systematic review methodologist across several databases and with different search term combinations to cover as many topics and conditions as possible. Moreover, grey literature, including clinical guidance, and funding body databases were also searched for research recommendations, planned and ongoing studies and funding calls, enabling the identification of gaps in knowledge. However, due to the rapid nature of REMs and certain streamlined process, such as study selection being conducted by one researcher, it is possible that available primary and secondary research studies were missed. Moreover, as women’s health is a broad topic, this REM had to be limited to certain topics, such as access to care, communication, mental health, endometriosis, and menopause. Thus, research gaps in other areas and health conditions, in which women might experience inequality, were not identified.

While performing the searches, different time limits were used for topics due to the varying volume of research published in certain areas. For example, time limit of 2012 was set for searches in access and communication, while limits of 2018 or 2021 were used for different endometriosis and menopause searches due to the high volume of research identified in these topics (detailed search strategy is presented in the Additional material). This step was necessary to make the REM manageable, although this might also mean that potentially relevant research published prior to the limits were missed. Furthermore, critical appraisal was not conducted to enable rapid production of this REM. While critical appraisal is an optional step in REMs, it is possible that the quality of included primary and secondary research studies might be low, which could also present further need for research.

## Data Availability

All data produced in the present work are contained in the manuscript

## Data Availability

The datasets generated during and/or analyzed during the current study are available from the corresponding author following reasonable request.

## 5. RAPID EVIDENCE MAP METHODS

### 5.1 Eligibility criteria

The eligibility criteria for the rapid evidence map, based on the Population, Phenomenon of Interest, Context, Study design (PiCoS) framework, are presented in Table 7.

**Table 7:**
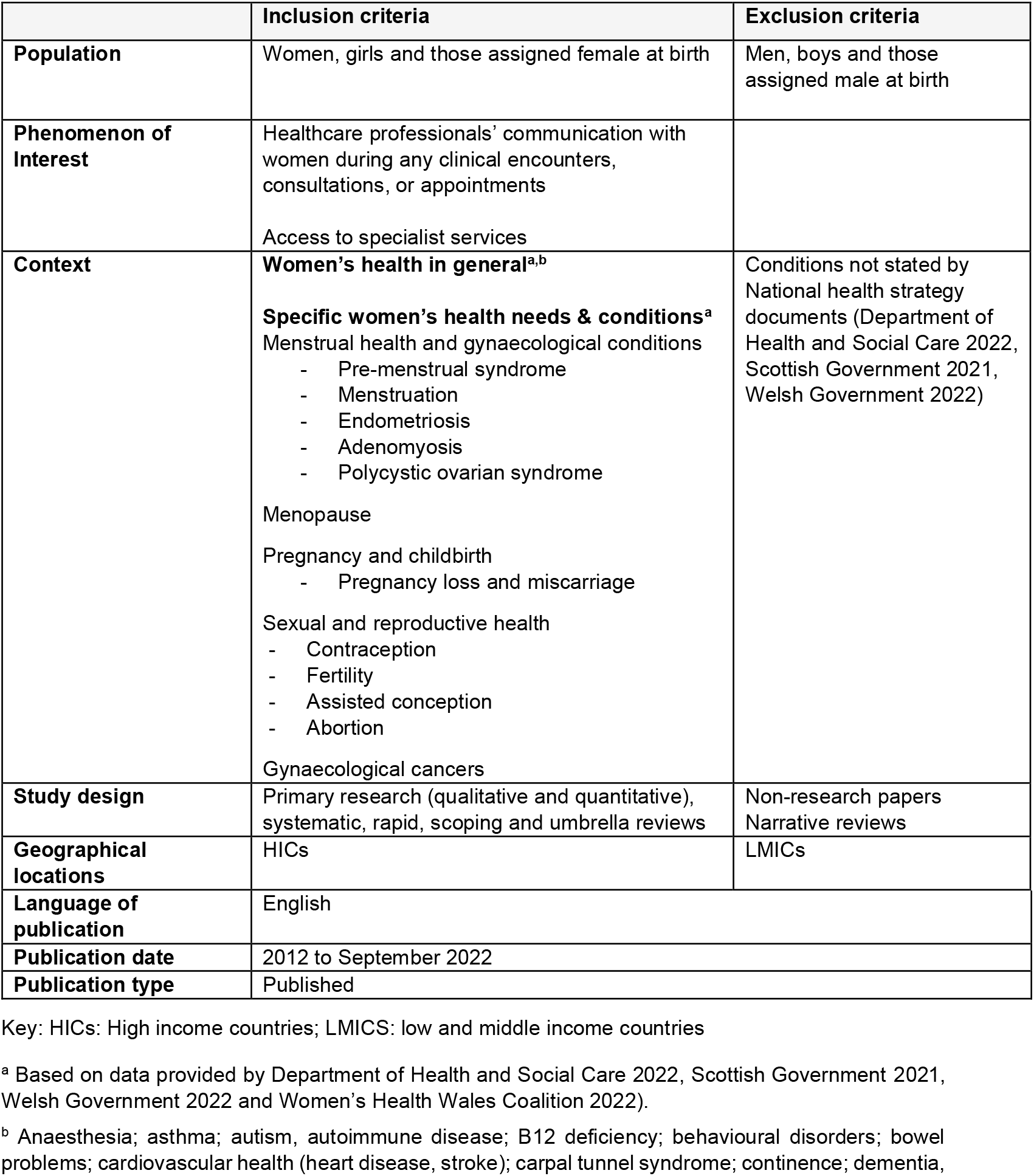
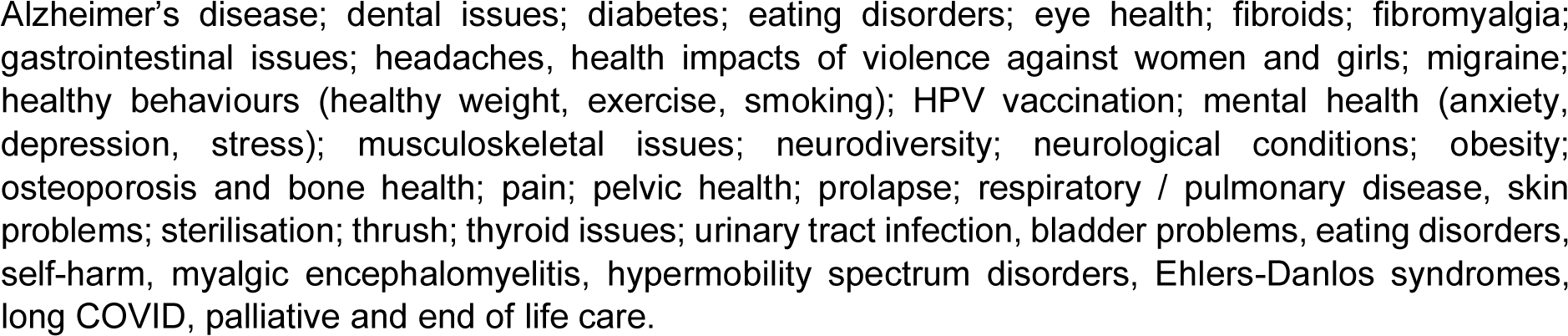
Eligibility Criteria.

|  | **Inclusion criteria** | **Exclusion criteria** |
| --- | --- | --- |
| **Population** | Women, girls and those assigned female at birth | Men, boys and those assigned male at birth |
| **Phenomenon of Interest** | Healthcare professionals' communication with women during any clinical encounters, consultations, or appointments<br>Access to specialist services |  |
| **Context** | **Women's health in general<sup>a,b</sup>**<br>**Specific women's health needs & conditions<sup>a</sup>**<br>Menstrual health and gynaecological conditions<br>- Pre-menstrual syndrome<br>- Menstruation<br>- Endometriosis<br>- Adenomyosis<br>- Polycystic ovarian syndrome<br>Menopause<br>Pregnancy and childbirth<br>- Pregnancy loss and miscarriage<br>Sexual and reproductive health<br>- Contraception<br>- Fertility<br>- Assisted conception<br>- Abortion<br>Gynaecological cancers | Conditions not stated by National health strategy documents (Department of Health and Social Care 2022, Scottish Government 2021, Welsh Government 2022) |
| **Study design** | Primary research (qualitative and quantitative), systematic, rapid, scoping and umbrella reviews | Non-research papers<br>Narrative reviews |
| **Geographical locations** | HICs | LMICs |
| **Language of publication** | English |  |
| **Publication date** | 2012 to September 2022 |  |
| **Publication type** | Published |  |

Key: HICs: High income countries; LMICS: low and middle income countries
<sup>a</sup> Based on data provided by Department of Health and Social Care 2022, Scottish Government 2021, Welsh Government 2022 and Women's Health Wales Coalition 2022).
<sup>b</sup> Anaesthesia; asthma; autism, autoimmune disease; B12 deficiency; behavioural disorders; bowel problems; cardiovascular health (heart disease, stroke); carpal tunnel syndrome; continence; dementia,
Alzheimer's disease; dental issues; diabetes; eating disorders; eye health; fibroids; fibromyalgia; gastrointestinal issues; headaches, health impacts of violence against women and girls; migraine; healthy behaviours (healthy weight, exercise, smoking); HPV vaccination; mental health (anxiety, depression, stress); musculoskeletal issues; neurodiversity; neurological conditions; obesity; osteoporosis and bone health; pain; pelvic health; prolapse; respiratory / pulmonary disease, skin problems; sterilisation; thrush; thyroid issues; urinary tract infection, bladder problems, eating disorders, self-harm, myalgic encephalomyelitis, hypermobility spectrum disorders, Ehlers-Danlos syndromes, long COVID, palliative and end of life care.

### 5.2 Evidence sources

Separate searches were conducted for different topics (such as communication, access, endometriosis, menopause, and mental health) and combination of topics. The searches were conducted across four databases MEDLINE (on the OVID platform), Embase (on the OVID platform), APA PSYCinfo (on the OVID platform) and CINAHL (on the EBSCO platform) with different date limits applied up to September 2022 for English language citations. Different date limits were necessary due to the high volume of records found in certain topic combinations. To make the rapid evidence map manageable, searches were limited for different dates (2012: communication, access; 2018: endometriosis and menopause research gaps, mental health; 2021: endometriosis and menopause reviews) depending on the volume of records found. The complete search strategy is presented in the [additional materials](http://www.primecentre.wales/resources/Supplementary/REM00045-Womens_Health_Additional_material_October-2022.pdf)

### 5.3 Search strategy

An initial key word search (within that title of a publication only) was undertaken on MEDLINE. The key words to be used were (women* OR woman* OR female AND health) AND access* OR encounter OR communication. Based on the initial search findings, an analysis of the text words contained in the title and abstract and of the index terms used to describe each article was conducted to inform the development of a search strategy. A series of search strategies were developed for different topics presented in this review, such as communication, access, endometriosis, menopause, and mental health. Detailed search strategies and results are presented in the [additional materials](http://www.primecentre.wales/resources/Supplementary/REM00045-Womens_Health_Additional_material_October-2022.pdf). The search strategies were tailored for each information source.

#### 5.3.1 Reference management

All reports retrieved from the database searches were imported or entered manually into reference management software EndNote™ and duplicates removed. At the end of this process the remaining reports were imported into web-based application Rayyan™.

### 5.4 Study selection process

One reviewer screened the reports using the information provided in the title and abstract using the web passed application Rayyan™. For reports that appear to meet the inclusion criteria, or in cases in which a definite decision cannot be made based on the title and/or abstract alone, the full texts of all reports were retrieved. The full texts were screened for inclusion by one reviewer. The flow of citations through each stage of the review process were displayed in a PRISMA flowchart.

### 5.5 Data extraction and coding/charting

The data extracted was extracted from the abstract only and included specific details about the populations, the focus of the research, study design and methodology, by one reviewer. No outcome data was extracted.

### 5.6 Assessment of methodological quality

An assessment of methodological quality was not conducted.

### 5.7 Data summary

The evidence was presented both narratively and in the form of tables and graphical evidence maps populated by information describing the number and types of studies and reviews by the women’s health conditions and also noted if any interventions had been conducted.

### 5.8 Further information available

- The Protocol is available on request
- [Search strategies](http://www.primecentre.wales/resources/Supplementary/REM00045-Womens_Health_Additional_material_October-2022.pdf)

## Abbreviations

Acronym: **Full Description**
BMA: British Medical Association
DHEA: Dehydroepiandrosterone
DHSC: Department of health and Social Care
FTWW: Fair Treatment for The Women of Wales
GSM: Genitourinary syndrome of menopause
HCP: Health care professionals
HCRW: Heath and Care Research Wales
HIC: High income countries
HRT: Hormone replacement therapy
HMB: Heavy menstrual bleeding
IPV: Intimate partner violence
NICE: National Institute of Clinical Excellence
NIHR: National Institute for Health and Care Research
OECD: Organisation for Economic Co-operation and Development
PCOS: Polycystic ovary syndrome
PMDD: Premenstrual dysphoric disorder
PMS: Premenstrual syndrome
REM: Rapid Evidence Map
VMS: Vasomotor symptoms
VTE: Venous thromboembolism
LNG-IUS: Levonorgestrel-releasing intra-uterine system

## 6.2 Acknowledgements

Michael Bowdery (Health and Care Research Wales); Richard Chivers (Women & Children’s Health Division); Lisa Daniels-Griffiths (Welsh Treasury), and Professor Jo Peden (Consultant in Public Health).

## 7. ABOUT THE WALES COVID-19 EVIDENCE CENTRE (WCEC)

The WCEC integrates with worldwide efforts to synthesise and mobilise knowledge from research.

We operate with a core team as part of Health and Care Research Wales, are hosted in the [Wales Centre for Primary and Emergency Care Research (PRIME),](http://www.primecentre.wales/) and are led by [Professor Adrian Edwards of Cardiff University](https://www.cardiff.ac.uk/people/view/123022-edwards-adrian).

The core team of the centre works closely with collaborating partners in [Health Technology Wales](https://www.cardiff.ac.uk/research/explore/research-units/wales-centre-for-evidence-based-care), Wales Centre for Evidence-Based Care, [Specialist Unit for Review Evidence centre](https://www.cardiff.ac.uk/specialist-unit-for-review-evidence), [SAIL Databank](https://healthandcareresearchwales.org/about-research-community/secure-anonymised-information-linkage-sail-databank), [Bangor Institute for Health & Medical Research/Health and Care Economics Cymru,](https://www.bangor.ac.uk/medical-health-sciences/research) and the [Public Health Wales Observatory](https://phw.nhs.wales/services-and-teams/observatory/).

Together we aim to provide around 50 reviews per year, answering the priority questions for policy and practice in Wales as we meet the demands of the pandemic and its impacts.

### Director

Professor Adrian Edwards

### Website

https://healthandcareresearchwales.org/about-research-community/wales-covid-19-evidence-centre

## 8. APPENDICES

### 8.1 Appendix 1: JLA Top 10 endometriosis research priority areas

1. Can a cure be developed for endometriosis?
  - What causes endometriosis?
  - What are the most effective ways of educating healthcare professionals throughout the healthcare system resulting in reduced time to diagnosis and improved treatment and care of women with endometriosis?
  - Is it possible to develop a non-invasive screening tool to aid the diagnosis of endometriosis?
  - What are the most effective ways of maximising and/or maintaining fertility in women with confirmed or suspected endometriosis?
  - How can the diagnosis of endometriosis be improved?
  - What is the most effective way of managing the emotional and/or psychological and/or fatigue impact of living with endometriosis (including medical, non-medical and self-management methods)?
  - What are the outcomes and/or success rates for surgical or medical treatments which aim to cure or treat endometriosis, rather than manage it?
  - What is the most effective way of stopping endometriosis progressing and/or spreading to other organs (e.g. after surgery)?
  - What are the most effective non-surgical ways of managing endometriosis-related pain and/or symptoms (medical/nonmedical)?

### 8.2 Appendix 2: NICE recommendations for research: endometriosis

1. Are pain management programmes a clinically and cost-effective intervention for women with endometriosis?
2. Is laparoscopic treatment (excision or ablation) of peritoneal disease in isolation effective for managing endometriosis-related pain?
3. Are specialist lifestyle interventions (diet and exercise) effective, compared with no specialist lifestyle interventions, for women with endometriosis? Studies should aim to provide evidence-based options to support self-management of endometriosis. This would improve the quality of life of women with endometriosis, enabling them to manage pain and fatigue, and reducing the negative impact on their career, relationships, sex lives, fertility, and physical and emotional wellbeing.
4. What information and support interventions are effective to help women with endometriosis deal with their symptoms and improve their quality of lives?

The direct effectiveness of different types or formats of information and support interventions on measurable outcomes such as health-related quality of life and level of function (for example, activities of daily living) have not been tested.

### 8.3 Appendix 3: NICE recommendations for research: menopause

#### Original guidance

- What is the safety and effectiveness of alternatives to systemic HRT as treatments for menopausal symptoms in women who have had treatment for breast cancer?
- What is the impact of systemic HRT usage in women with a previous diagnosis of breast cancer for the risk of breast cancer reoccurrence, mortality or tumour aggression?
- How does the preparation of HRT affect the risk of venous thromboembolism (VTE)?
- What is the difference in the risk of breast cancer in menopausal women on HRT with progesterone, progestogen or selective oestrogen receptor modulators?
- What is the impact of oestradiol in combination with the levonorgestrel-releasing intra- uterine system (LNG-IUS) on the risk of breast cancer and VTE?
- What are the effects of early HRT use on the risk of dementia?
- .What are the main clinical manifestations of premature ovarian insufficiency and the short- and long-term impact of the most common therapeutic interventions?

#### Updated guidance

The following areas have been identified for inclusion in the scope:

- Managing menopausal symptoms.
- Cognitive behavioural therapy to manage symptoms associated with the menopause.
- Interventions to manage genitourinary symptoms associated with the menopause.
- Effects of hormone replacement therapy on overall health outcomes.

The surveillance and scoping process did not identify any substantive new evidence on using testosterone beyond the current recommendations in the NICE guideline for using testosterone for altered sexual function. NICE discussed the need for evidence in this area with the NIHR who have agreed to scope new research.

### 8.4 Appendix 4: The BMS recommendations for research: menopause

- Vaginal DHEA use in cancer survivors.
- The effects of acupuncture on VMS before it can be considered a more effective therapy than placebo.
- Efficacy and safety of oral DHEA
- DHEA pessaries have recently been licensed for the treatment of vulvovaginal atrophy and may have some benefits for low libido. However, this requires further evaluation in adequately powered randomised studies.
- St John’s wort and some isoflavone preparations may be effective for VMS but more research is required to confirm efficacy.
- Head-to-head comparisons of safety and efficacy of vaginal DHEA to topical vaginal oestrogens are not available and further research is required to assess this.

### 8.5 Appendix 5: NICE recommendations for research: adenomyosis

- The effects of uterine artery embolisation compared with other procedures to treat adenomyosis, particularly for patients wishing to maintain or improve their fertility.

### 8.6 Appendix 6: NICE recommendations for research: uterine fibroids

- No recommendations for research within the guidance.

### 8.7 Appendix 7: NICE recommendations for research: heavy menstrual bleeding

- Hysteroscopy compared with ultrasound or empiric pharmacological treatment in the diagnosis and management of heavy menstrual bleeding (HMB).
- Effectiveness of the progestogen-only pill, injectable progestogens, or progestogen implants in alleviating HMB.
- Long-term outcomes of pharmacological and uterine-sparing surgical treatments for HMB associated with adenomyosis.
- Hysteroscopic removal of submucosal fibroids compared with other uterine-sparing treatments for HMB.
- Are outcomes after second-generation endometrial ablation for women with HMB associated with myometrial pathology (adenomyosis and/or uterine fibroids) equivalent to those for women without myometrial pathology?

### 8.8 Appendix 8: The National Association for Premenstrual Syndrome: Guidelines on Premenstrual Syndrome

This guidance did not include specific recommendations for research into premenstrual syndrome (PMS) but did highlight some areas where there is insufficient data.

- Insufficient evidence of efficacy is available to give a recommendation for using Vitamin B6 in the treatment of PMS.
- Given that calcium and Vitamin D may also reduce the risk of osteoporosis and some cancers, clinicians may consider recommending these nutrients even for women with PMS, but more data are required to determine efficacy and to optimise regimens.
- More data are required before a clear recommendation can be made for isoflavone usage, but preliminary data are encouraging.
- Agnus Castus is the best researched complimentary therapy for PMS, but a lack of standardised quality controlled preparations is a problem.
- Initial data appear encouraging but larger studies are required before St John’s Wort can be recommended for use in PMS.
- There are insufficient data to recommend the routine use of progestogens or natural progesterone in the treatment of PMS.

### 8.9 Appendix 9: NICE recommendations for research: Fertility problems (PCOS)

- No recommendations for research included within the guidance.

